# A call for governments to pause Twitter censorship: a cross-sectional study using Twitter data as social-spatial sensors of COVID-19/SARS-CoV-2 research diffusion

**DOI:** 10.1101/2020.05.27.20114983

**Authors:** Vanash M. Patel, Robin Haunschild, Lutz Bornmann, George Garas

## Abstract

**Objectives:** To determine whether Twitter data can be used as social-spatial sensors to show how research on COVID-19/SARS-CoV-2 diffuses through the population to reach the people that are especially affected by the disease.

**Design:** Cross-sectional bibliometric analysis conducted between 23^rd^ March and 14^th^ April 2020.

**Setting:** Three sources of data were used in the analysis: (1) deaths per number of population for COVID-19/SARS-CoV-2 retrieved from Coronavirus Resource Center at John Hopkins University and Worldometer, (2) publications related to COVID-19/SARS-CoV-2 retrieved from WHO COVID-19 database of global publications, and (3) tweets of these publications retrieved from Altmetric.com and Twitter.

**Main Outcome(s) and Measure(s):** To map Twitter activity against number of publications and deaths per number of population worldwide and in the USA states. To determine the relationship between number of tweets as dependent variable and deaths per number of population and number of publications as independent variables.

**Results:** Deaths per one hundred thousand population for countries ranged from 0 to 104, and deaths per one million population for USA states ranged from 2 to 513. Total number of publications used in the analysis was 1761, and total number of tweets used in the analysis was 751,068. Mapping of worldwide data illustrated that high Twitter activity was related to high numbers of COVID-19/SARS-CoV-2 deaths, with tweets inversely weighted with number of publications. Poisson regression models of worldwide data showed a positive correlation between the national deaths per number of population and tweets when holding the country’s number of publications constant (coefficient 0.0285, S.E. 0.0003, p<0.001). Conversely, this relationship was negatively correlated in USA states (coefficient –0.0013, S.E. 0.0001, p<0.001).

**Conclusions:** This study shows that Twitter can play a crucial role in the rapid research response during the COVID-19/SARS-CoV-2 global pandemic, especially to spread research with prompt public scrutiny. Governments are urged to pause censorship of social media platforms during these unprecedented times to support the scientific community’s fight against COVID-19/SARS-CoV-2.

**SUMMARY BOX:** *What is already known on this topic:*

- Twitter is progressively being used by researchers to share information and knowledge transfer.
- Tweets can be used as ‘social sensors’, which is the concept of transforming a physical sensor in the real world through social media analysis.
- Previous studies have shown that social sensors can provide insight into major social and physical events.

*What this study adds:*

- Using Twitter data used as social-spatial sensors, we demonstrated that Twitter activity was significantly positively correlated to the numbers of COVID-19/SARS-CoV-2 deaths, when holding the country’s number of publications constant.
- Twitter can play a crucial role in the rapid research response during the COVID-19/SARS-CoV-2 global pandemic.

## INTRODUCTION

Twitter is a social network created in 2006, that brings together hundreds of millions of users around its minimalist concept of microblogging, allowing users to post and interact with messages known as ‘tweets”.(1) Twitter has short delays in reflecting what its users perceive, and its principle of “following” users without obligatory reciprocity, together with a very open application programming interface, make it an ideal medium for the study of online behaviour.(2) Tweets can be used as ‘social sensors’, which is the concept of transforming a physical sensor in the real world through social media analysis. Tweets can be regarded as sensory information and Twitter users as sensors. Studies have demonstrated that tweets analysed as social sensors can provide insight into major social and physical events like earthquakes (3), sporting events (4), celebrity deaths (5), and presidential elections.(6) Twitter data contain location information which can be converted into geo-coordinates and be spatially mapped. In this way tweets can be used as social-spatial sensors to demonstrate how research diffuses within a population.(7)

Researchers are increasingly using Twitter as a communication platform, and tweets often contain citations to scientific papers.(8) Twitter citations can form part of a rapid dialogue between users which may express and transmit academic impact and support traditional citation analysis. Twitter citations are defined ‘as direct or indirect links from a tweet to a peer-reviewed scholarly article online’ (3, 8), and reflect a broader discussion crossing traditional disciplinary boundaries, as well as representing ‘attention, popularity or visibility’ rather than influence.(9) Coronavirus disease 2019 (COVID-19) is a novel infectious disease caused by severe acute respiratory syndrome coronavirus 2 (SARS-CoV-2). The World Health Organization (WHO) declared the 2019–20 coronavirus outbreak a Public Health Emergency of International Concern (PHEIC)(10) on 30 January 2020 and a pandemic on 11 March 2020.(11)

We use Twitter data as social-spatial sensors to demonstrate how research on COVID-19/SARS-CoV-2 diffuses through the population and to investigate whether research reaches the people that are especially affected by the disease.

## METHODS

### Dataset used

We used three sources of data in this study: (1) deaths per number of population for COVID-19/SARS-CoV-2, (2) publications related to COVID-19/SARS-CoV-2, and (3) tweets of these publications. All data was retrieved and analysed between 23^rd^ March and 14^th^ April 2020.

### Deaths

We used deaths per number of population as a measure of severity of the outbreak of the virus in countries and USA states. We used deaths per one hundred thousand population for country specific data, which was retrieved from Coronavirus Resource Center at John Hopkins University.(12) We used deaths per one million population for US state specific data, which was retrieved from Worldometer, a provider of global COVID-19 statistics trusted by institutions such as the United Kingdom government and The Center for Systems Science and Engineering at Johns Hopkins University.(13)

### Publications

We used the WHO COVID-19 database of global publications, which integrates the latest international multilingual scientific findings and knowledge on COVID-19 from searches of bibliographic databases, hand searching, and the addition of other expert-referred scientific articles (see supplementary material).(14)

### Tweets

We used the Altmetric.com application programming interface to extract tweet identifiers for any tweets which mentioned any of the publications on Twitter (see supplementary material).(15)

### Statistical analysis

We used several Stata commands in this study.(16–18) The most important Stata commands were `shp2dta` (19) and `spmap` (20) to produce the Twitter maps. We additionally calculated Poisson regression models with number of tweets as dependent variable and COVID-19/SARSCoV-2 deaths per number of population and number of publications as independent variables. We included another binary independent variable reflecting national censorship of Twitter in Iran and China (1 = national censorship). With count variables as dependent variables, Poisson regression models are indicated.(21, 22) We focus on percentage changes in expected counts in the interpretation of the models.(23) These percentages can be interpreted as follows: for a standard deviation increase in the death rate per population in a country (or US state), the increases in the expected tweet number in that country (or US state), holding the country’s (or US state’s) number of publications constant.

## RESULTS

The deaths per one hundred thousand population for countries ranged from 0 (Ethiopia) to 104 (San Marino). The deaths per one million population for USA states ranged from 2 (Wyoming) to 513 (New York). The total number of publications that were used in the analysis was 1761, and the total number of tweets that were used in the analysis was 751,068 (see supplementary material). **Figure 1**, which shows a histogram of tweets per day since 17^th^ November 2020 (date of the first known case of COVID-19/SARS-CoV-2), demonstrates exponential growth in twitter activity in March 2020.

**Figure 1:**
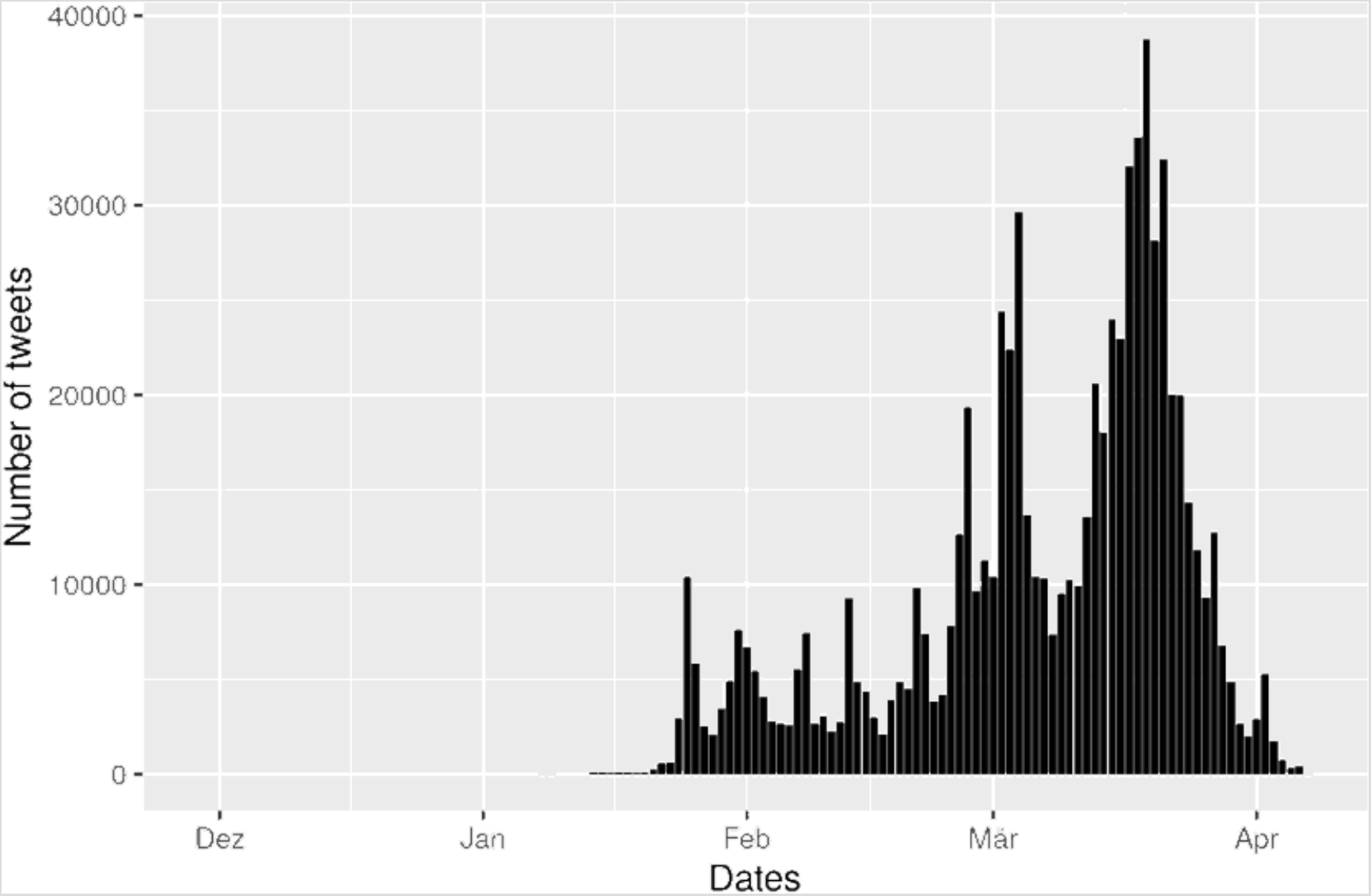
Tweet timeline showing number of tweets per day since 17^th^ November 2020

### Mapping worldwide data

**Figure 2** shows worldwide Twitter activity referring to publications dealing with COVID-19/SARS-CoV-2. The underlying blue-colored scheme visualizes national deaths per number of population. The map is intended to show whether COVID-19/SARS-CoV-2 research reaches regions with many COVID-19/SARS-CoV-2 deaths: does the number of COVID-19/SARSCoV-2 cases correlate with the number of tweets on COVID-19/SARS-CoV-2 publications?

**Figure 2:**
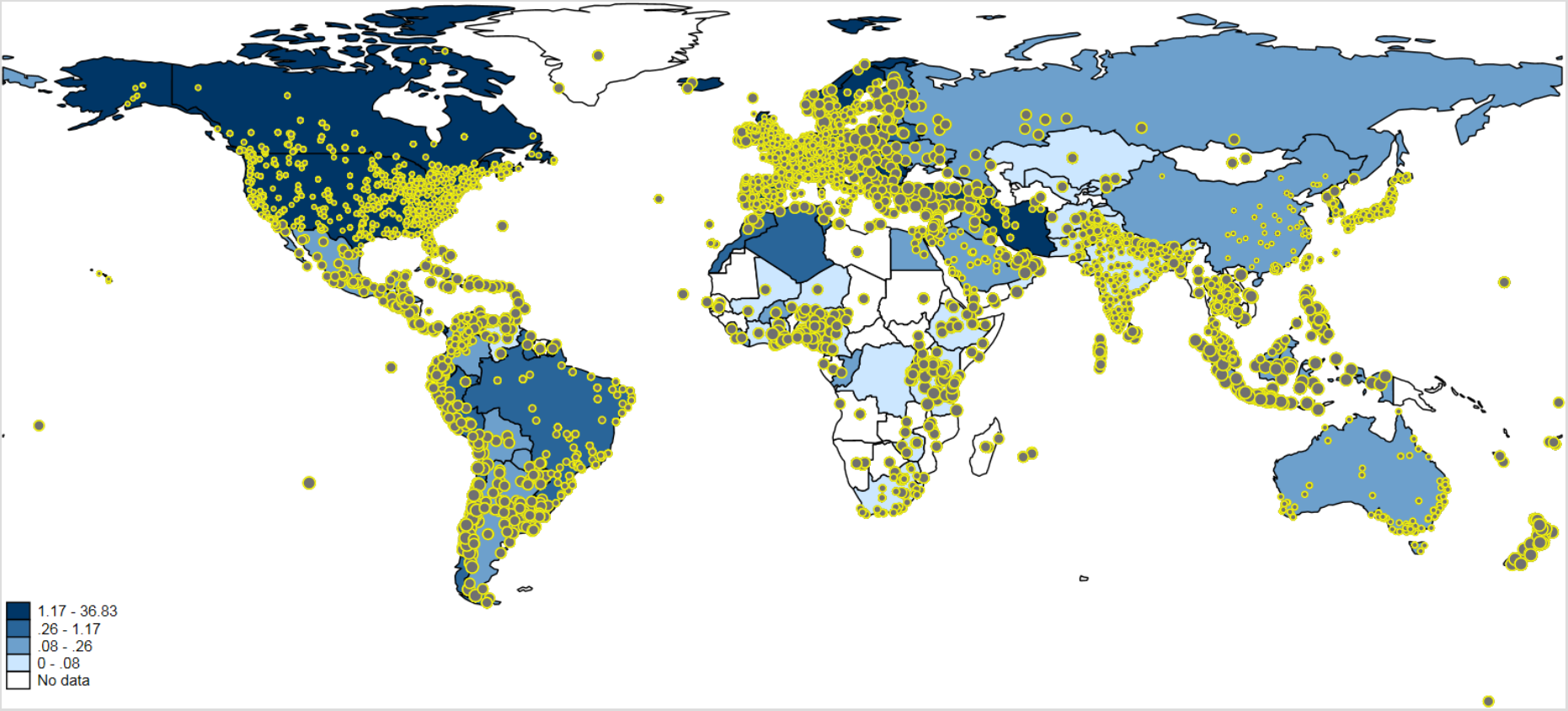
Tweeting on publications dealing with COVID-19/SARS-CoV-2 worldwide. Each tweet is inversely weighted with the number of publications published by authors in the corresponding country: the larger the dots, the smaller the research activity. The countries are colored according to the national deaths per one hundred thousand population. For some countries, e.g. Greenland, no data are available. Countries such as China and Iran block internet access to Twitter or its content.(12)

One of the problems with Twitter data in the context of this study is that Twitter activity is generally high where more research is done (e.g., Western Europe or the Boston region in Figure 2). Since this is not the activity which we intended to measure, we inversely weighted the size of each tweet on the map by the number of publications in that country [i.e., 1/log(number of publications)]. Thus, the more publications’ authors are located in a country, the smaller the size of the tweet dot is (see here 24, 25). We assume that large dots reflect tweets of people not doing research or not being a publisher/publishing organization (but might be personally confronted with COVID-19/SARS-CoV-2).

The map in **Figure 2** might show the expected result that high Twitter activity is related to high numbers of COVID-19/SARS-CoV-2 deaths. However, it is not completely clear whether this conclusion can be drawn, since there are several countries with high Twitter activity and high publication output (e.g., Western Europe and the Boston region). For some regions on the map, the extent of Twitter activity is difficult to interpret since tweet dots might overlap (especially those with larger sizes). To have a conclusive answer on the relation between Twitter activity and publication output, we additionally calculated Poisson regression models with number of tweets as dependent variable and deaths per number of population and number of publications as independent variables.

The results are shown in **Table 1**. The coefficients of deaths per number of population and number of publications are statistically significant. The percentage changes in expected counts reveal that deaths per number of population and Twitter activities are related in fact: for a standard deviation increase in the national deaths per number of population, the expected number of tweets in that country increases by 19.7%, holding the country’s number of publications constant. The results in. further show that the influence of the number of publications is significantly higher than that of deaths per number of population.

**Table 1.** Coefficients of a Poisson regression model with number of tweets as dependent variable (n=111 countries)

| Independent variable | Coefficient | Standard error | Percentage change in expected count |
| --- | --- | --- | --- |
| Deaths per number of population | 0.0285*** | 0.0003 | 19.7 |
| Number of publications | 0.0152*** | $2.82 \times 10^{-5}$ | 155.5 |
| National censorship of Twitter | -8.6662*** | 0.0376 |  |
| Constant | 6.3950*** | 0.0040 |  |

Notes. \*\*\* $p < .001$

### Mapping United States of America (USA) data

We did not only use the Twitter data as social-spatial sensors to investigate global trends, but also within a single country. **Figure 3** shows publication-based Twitter activity dealing with COVID-19/SARS-CoV-2 in the USA. The blue-colored scheme presents the deaths in the USA states per one million population. The map might show that the deaths in the USA states are in fact related to the number of tweets on COVID-19/SARS-CoV-2 publications. However, there are several USA states with high Twitter activity and high publication output (e.g., the Boston region).

**Figure 3:**
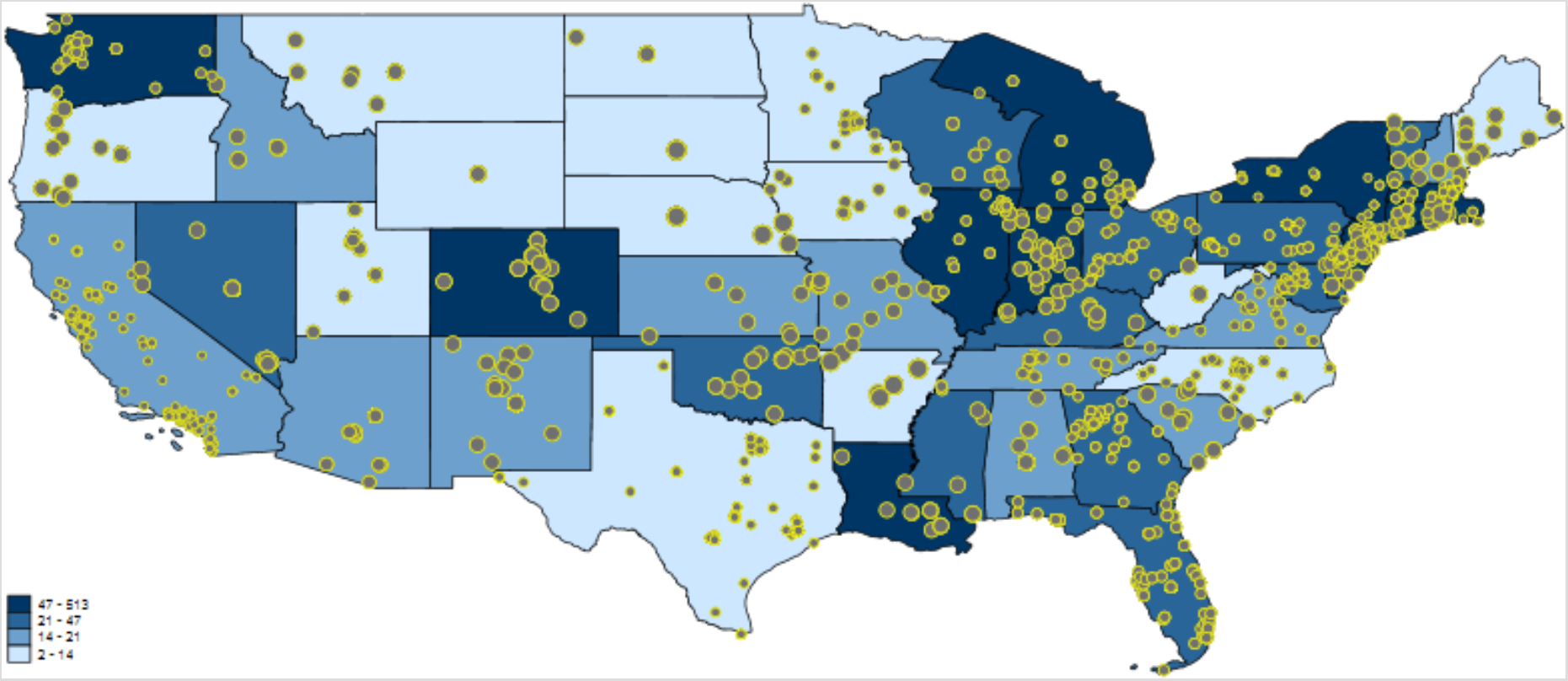
Tweeting on publications dealing with COVID-19/SARS-CoV-2 in the USA. Each tweet is inversely weighted with the number of publications published by authors in the corresponding USA state: the larger the dots, the smaller the research activity. The USA states are colored according to their deaths per one million population.(13)

We calculated Poisson regression models with deaths per number of population and number of publications as independent variables and number of tweets as dependent variable. **Table 2** reports the results. The results are based on 49 USA states (out of 52) since only USA states with at least one tweet were considered. The percentage changes in expected counts in Table 2 point out that deaths per number of population and Twitter activities are negatively correlated: for a standard deviation increase in the deaths per number of population of a USA state, the expected number of tweets in that state decreases by 10.6%, holding the USA state’s number of publications constant. The results in Table 2 further show that the influence of the number of publications is significantly greater than that of the deaths per number of population (and positive). In the USA states, there is a strong dependency of Twitter data on the number of publications. **Figure S1** demonstrates that at the time of the analysis the USA was an outlier because of lower national deaths per number of population and higher numbers of publications and tweets, when compared to other countries that were significantly impacted by COVID-19/SARS-CoV-2 (e.g., UK, France, Spain, and Italy).

**Table 2.** Coefficients of Poisson regression model with number of tweets as dependent variable (n=49 USA states)

| Independent variable | Coefficient | Standard error | Percentage change in expected count |
| --- | --- | --- | --- |
| Deaths per number of population | -0.0013*** | 0.0001 | -10.6 |
| Number of publications | 0.0744*** | 0.0005 | 105.5 |
| Constant | 5.1767*** | 0.0114 |  |

Note. \*\*\* $p < .001$

## DISCUSSION

This study demonstrates that Twitter data can be used as social-spatial sensors to monitor research diffusion in a global pandemic using COVID-19/SARS-CoV-2 as an example. Our results suggest that novel research on COVID-19/SARS-CoV-2 publicised through Twitter reaches populations that are concerned about the disease.

Social media can be an effective tool for broadcasting research both within and beyond the academic community.(26) Twitter is one of the best social media platforms for sharing scientific research and knowledge because it allows users to post links of recent publications, write a short statement about the research topic and tag keywords with hashtags, so that people who are interested in the research are more likely to see the post.(27) As well as promoting scientific research, Twitter and other social media platforms can scrutinise research in public, often within hours rather than years, unearthing poor quality inaccurate work.(27) Governments and research institutions worldwide support a rapid research response to improve understanding of COVID-19/SARS-CoV-2, including the development and testing of therapies and vaccines.(28) Our study shows that Twitter can play a vital role in the rapid research response, especially in disseminating research with swift peer review. Our study shows exponential use of Twitter as the intensity of the outbreak has increased. Over 80% of publications extracted from the WHO COIVD-19 database have been cited on Twitter, which is nearly seven times higher than previous studies analysing Twitter data in biomedical sciences.(29) Each COVID-19 publication has been tweeted on average 425 times, which is significantly higher than our previous work analyzing Twitter activity of single infectious diseases (on average publications related to Human Immunodeficiency Virus (HIV) were tweeted 7 times, tuberculosis were tweeted 8 times, and malaria tweeted 9 times).(7)

Countries such as China and Iran have blocked Twitter, as well as other social media platforms.(30) This is reflected in our mapping of worldwide tweet data related to research on COVID-19/SARS-CoV-2 (Figure 2). The COVID-19/SARS-CoV-2 originated from Wuhan in China’s Hubei province, which quickly became the epicentre for China’s outbreak, followed by a new epicentre in Iran. Both countries have seen a rapid rise in scientific output over the last two decades and their research (31), coupled with thousands of reported cases on COVID-19/SARS-CoV-2 has led them to a better understanding of the novel, fast moving virus. However, censorship of social media may have stifled research dissemination and more importantly avoids swift public scrutiny. This may adversely affect the global fight against the disease. Our study suggests that governments should consider relaxing censorship of social media at times of global crisis, such as the COVID-19/SARS-CoV-2 pandemic. Moreover, allowing researchers greater access to platforms such as Twitter during a global pandemic can aid the scientific community’s fight against misinformation and pseudoscience.(32)

The USA appears to be an outlier in the worldwide data and the country-specific data shows that the USA has a different relationship between tweets and deaths, both of which may be due to the pandemic reaching the USA later than most other countries in the Northern Hemisphere. Another explanation is the difference in geographical clusters of COVID-19/SARS-CoV-2 and research productivity in the USA. On one hand, New York state became the global epicentre of the pandemic after the virus spread through Europe, and over a third of USA COVID-19/SARSCoV-2 deaths have occurred in New York state, with the majority in New York City.(13) On the other hand, the USA is the most prolific publisher of high-quality science in the world, but the top-performing institutes are concentrated in Massachusetts, California and Maryland.(33)

Before concluding, it is important to consider the limitations of this study. We have analysed tweets mentioning publications in a quantitative manner which does not account for the association of the tweet with the publication (i.e. a tweet may reference a valid study but claim it to be ‘fake news’ or have another negative overtone). We have not performed any thematic analysis of the tweets in terms of their content (e.g., are tweets referring to testing for COVID-19/SARS-CoV-2, therapies, or vaccines), quality (e.g., whether tweets are referring to randomised controlled trials or letters), or who tweeted these (e.g., individual researchers, members of the public, universities or pharmaceutical industries). Moreover, no distinction was made between tweets and retweets (of original tweets), which raises the question whether a different handling of retweets could yield different results. These are interesting questions which might be an interesting topic of further research.

Despite these limitations, our study has a number of strengths. We have used an evidence-based and robust methodology (see supplementary material) to clean and analyse data, as well as extracting data from several well-established databases containing real world evidence updated in real time.(12–15) Our study comes at a very critical point in time, when a rapid research response is vital to develop therapies and vaccines to slow the COVID-19/SARS-CoV-2 pandemic and lessen the damage caused by the disease. Our study utilising Twitter data as social-spatial sensors can serve as proof-of-concept for future studies on Twitter and the evolving pandemic.

## CONCLUSION

COVID-19/SARS-CoV-2 began as a cluster of cases of pneumonia in Wuhan, Hubei Province, but the outbreak quickly progressed from an PHEIC to a pandemic, which highlights the dynamic process of the spread of an infectious disease.(10, 11) Our study has simply investigated a snapshot of the relationship between this pandemic, research outputs, and Twitter activity, and demonstrates the importance of how social media platforms can be crucial to spread research with rapid scrutiny, which may also impede the degree of misinformation. We urge governments to pause censorship of social media platforms such as Twitter during these unprecedented times to support the scientific community’s battle against COVID-19/SARSCoV-2.

## Data Availability

The full data set and the statistical code can be obtained, upon request, from the corresponding author.

## ACKNOWLEDGEMENTS

Meta-data for publications were downloaded via the Dimensions API. Twitter data were retrieved from the Altmetric.com API. Tweets with their location information were retrieved from the Twitter API. The authors thank Rodrigo Costas (CWTS) and Stacy Konkiel (Altmetric.com) for helpful discussions regarding the analysis of location information of Twitter users.

## TRANSPARENCY DECLARATION

The lead author affirms that the manuscript is an honest, accurate, and transparent account of the study being reported. No important aspects of the study have been omitted, and any discrepancies from the study as planned have been explained.

## ETHICS COMMITTEE APPROVAL

Not applicable

